# The impact of demographic factors on the accumulated number of COVID-19 cases per capita in Europe and the regions of Ukraine in the summer of 2021

**DOI:** 10.1101/2021.07.04.21259980

**Authors:** Igor Nesteruk, Oleksii Rodionov

**Affiliations:** Institute of Hydromechanics, National Academy of Sciences of Ukraine, Kyiv, Ukraine; Igor Sikorsky Kyiv Polytechnic Institute, Kyiv, Ukraine; Private consulting office, Kyiv, Ukraine

**Keywords:** COVID-19 pandemic, epidemic dynamics in Europe, epidemic dynamics in Ukraine, mathematical modeling of infection diseases, statistical methods

## Abstract

The accumulated number of COVID-19 cases per capita is an important characteristic of the pandemic dynamics that may also indicate the effectiveness of quarantine, testing and vaccination. As this value increases monotonically over time, the end of June 2021 was chosen, when the growth rate in Ukraine and the vast majority of European countries was small. This allowed us to draw some intermediate conclusions about the influence of the volume of population, its density, and the level of urbanization on the accumulated number of laboratory-confirmed cases per capita in European countries and regions of Ukraine. A simple analysis showed that the number of cases per capita does not depend on these demographic factors, although it may differ by about 4 times for different regions of Ukraine and more than 9 times for different European countries. The number of COVID-19 per capita registered in Ukraine is comparable with the same characteristic in other European countries but much higher than in China, South Korea and Japan.

## Introduction

The accumulated number of COVID-19 cases per capita (CC) may indicate the effectiveness of quarantine, testing, vaccination, and also characterizes the virulence of coronavirus strains circulating in a particular region. The CC values increase monotonically over time, so it is important to fix the appropriate time and compare these values for different countries and regions. In particular, in this study we take the end of June 2021, when the CC growth rate in Ukraine and the vast majority of European countries was small. The CC numbers are regularly reported by World Health Organization, [1] and COVID-19 Data Repository by the Center for Systems Science and Engineering (CSSE) at Johns Hopkins University (JHU), [2].

The impact of some eco-demographic factors on the COVID-19 pandemic dynamics was studied in [3-19]. The influence of the population volume *N*_*pop*_ on the final sizes *V*_∞_ of the first pandemic waves in different countries and regions was studied in [19] and compared with the real CC values at fixed moments of time. In particular, relative final size of the first epidemic wave 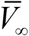 was approximated by following equations:

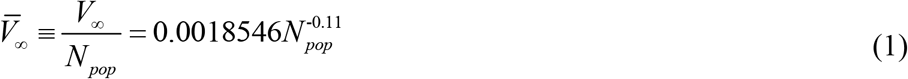

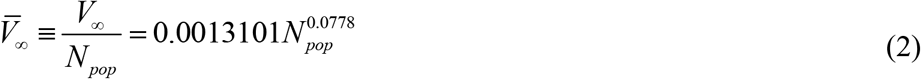

with the use results of SIR simulations for n=13 countries and regions and the results for n=11 (without mainland China and South Korea), respectively.

In this paper we will study the influence of the volume of population *N*_*pop*_, its density, and the level of urbanization *N*_*ubr*_/*N*_*pop*_ (*N*_*ubr*_ is the number of people living in cities) on the accumulated number of laboratory-confirmed cases per capita in European countries and regions of Ukraine.

### Data

We will use the data set regarding the numbers of laboratory-confirmed COVID-19 cases in the regions of Ukraine accumulated at the time June 27, 2021 and registered by national statistics, [20]. The corresponding CC numbers (per 100 persons of population) and demographic data sets for Ukrainian regions [21] are shown in Table 1. As the information from the regions of Ukraine fully or partially occupied by the Russian Federation is inaccurate, we excluded from consideration Donetsk and Luhansk regions, Crimea and Sevastopol. It can be seen that CC values vary from 2.25 (Kirovohrad oblast) to 8.84 (Chernivtsi oblast). Rather high CC values were registered in Zhytomyr, Khmelnytskyi, Kyiv (oblast and city), and Sumy regions that are not in the west of Ukraine with traditionally close ties with the EU countries.

**Table 1.** Demographic characteristics and the accumulated number of laboratory-confirmed COVID-19 cases in the regions of Ukraine as of June 27, 2021.

| **Oblast (region)** | **Population,**<br>**$N_{pop}$,**<br>**[21]** | **Urban**<br>**population,**<br>**$N_{ubr}$,**<br>**[21]** | **Population**<br>**density (number**<br>**of people per**<br>**square**<br>**kilometer), [21]** | **Accumulated**<br>**number of**<br>**laboratory-**<br>**confirmed cases**<br>**per hundred,**<br>**[20]** |
| --- | --- | --- | --- | --- |
| Vinnitsia | 1 545 416 | 799 385 | 58.29 | 4.67 |
| Volyn | 1 031 421 | 539 179 | 51.2 | 6.06 |
| Dnipropetrovsk | 3 176 648 | 2 668 744 | 99.54 | 4.33 |
| Zhytomyr | 1 208 212 | 716 457 | 40.5 | 7.43 |
| Zakarpattia | 1 253 791 | 465 904 | 98.13 | 4.98 |
| Zaporizhzhia | 1 687 401 | 1 306 231 | 62.08 | 6.30 |
| Ivano-Frankivsk | 1 368 097 | 606 764 | 98.23 | 6.38 |
| Kyiv (oblast) | 1 781 044 | 1 105 383 | 63.31 | 7.12 |
| Kirovohrad | 933 109 | 591 944 | 37.95 | 2.25 |
| Lviv | 2 512 084 | 1 534 040 | 115.06 | 5.52 |
| Mykolaiv | 1 119 862 | 768 022 | 45.53 | 6.34 |
| Odesa | 2 377 230 | 1 597 062 | 71.37 | 5.95 |
| Poltava | 1 386 978 | 867 201 | 48.25 | 5.70 |
| Rivne | 1 152 961 | 548 088 | 57.51 | 6.93 |
| Sumy | 1 068 247 | 741 430 | 44.82 | 7.43 |
| Ternopil | 1 038 695 | 473 727 | 75.14 | 6.82 |
| Kharkiv | 2 658 461 | 2 158 121 | 84.62 | 5.66 |
| Kherson | 1 027 913 | 631 317 | 36.12 | 3.53 |
| Khmelnyskyi | 1 254 702 | 720 752 | 60.82 | 7.18 |
| Cherkasy | 1 192 137 | 678 682 | 57.04 | 6.97 |
| Chernivtsi | 901 632 | 390 551 | 111.35 | 8.94 |
| Chernihiv | 991 294 | 649 063 | 31.11 | 5.92 |
| Kyiv (city) | 2 967 360 | 2 967 360 | 3536.78 | 7.22 |

The CC figures (per 1,000,000 persons of population) accumulated at the time June 28, 2021 and registered by JHU, [2] are shown in Table 2, which contains also the demographic data sets for European countries taken from [22-24]. The highest CC levels were registered in Andorra - 18%, Montenegro – 16%, Czech Republic -15.5%, San Marino -15%, Slovenia -12.4%. The populations of these countries are not very high as well as populations of Iceland and Finland where the lowest CC values were registered (1.9% and 1.7%, respectively). More than a tenfold difference in values raises a reasonable question about its causes. They may be related to the population density or the level of urbanization. A possible relationship with these factors will be explored further.

**Table 2.** Demographic characteristics and the accumulated number of laboratory-confirmed COVID-19 cases in European countries as of June 28, 2021.

| Country | Population,<br>[22] | Urbanization<br>$N_{ubr}/N_{pop}$, %, [23] | Population<br>density (per<br>quare km), [24] | Total cases per<br>million, [2] |
| --- | --- | --- | --- | --- |
| Monaco | 38682 | 100 | 15,47 | 65615.126 |
| Holy See (Vatican City) | 801 | 100 | 2,273 | 33374.536 |
| Malta | 514564 | 95 | 1,447 | 69330.229 |
| San Marino | 33785 | 97 | 561 | 150008.84 |
| Netherlands | 17059560 | 91 | 416 | 99886.646 |
| Belgium | 11482178 | 98 | 384 | 93486.963 |
| United Kingdom | 67141684 | 83 | 270 | 70284.988 |
| Liechtenstein | 37910 | 14 | 245 | 79555.288 |
| Luxembourg | 604245 | 91 | 243 | 112850.972 |
| Germany | 83124418 | 77 | 225 | 44576.917 |
| Italy | 60627291 | 70 | 207 | 70432.141 |
| Switzerland | 8525611 | 74 | 204 | 81198.962 |
| Andorra | 77006 | 88 | 183 | 179667.379 |
| Denmark | 5752126 | 88 | 136 | 50743.387 |
| Czech Republic | 10665677 | 74 | 136 | 155658.773 |
| Poland | 37921592 | 60 | 122 | 76088.436 |
| Portugal | 10256193 | 65 | 112 | 85856.051 |
| Slovakia | 5453014 | 54 | 111 | 71720.074 |
| Albania | 2882740 | 60 | 107 | 46046.633 |
| Austria | 8891388 | 66 | 106 | 72206.875 |
| France | 64990511 | 80 | 105 | 86325.089 |
| Hungary | 9707499 | 71 | 105 | 83645.21 |
| Turkey | 82340088 | 75 | 105 | 64196.94 |
| Slovenia | 2077837 | 55 | 104 | 123742.383 |
| Moldova | 4051944 | 43 | 99 | 63613.375 |
| Spain | 46692858 | 80 | 99 | 81117.733 |
| Serbia | 8802754 | 56 | 91 | 105279.579 |
| Romania | 19506114 | 54 | 89 | 56174.491 |
| North Macedonia | 2082957 | 58 | 83 | 74722.806 |
| Greece | 10522246 | 79 | 80 | 40416.745 |
| Bosnia and Herzegovina | 3323925 | 48 | 75 | 62481.731 |
| Croatia | 4156405 | 57 | 75 | 87610.845 |
| Ireland | 4818690 | 63 | 74 | 55002.07 |
| Ukraine | 44246156 | 69 | 73 | 52556.15 |
| Bulgaria | 7051608 | 75 | 63 | 60682.066 |
| Belarus | 9452617 | 78 | 46 | 44001.045 |
| Montenegro | 627809 | 67 | 44 | 159544.758 |
| Lithuania | 2801264 | 68 | 42 | 102372.965 |
| Latvia | 1928459 | 68 | 29 | 72759.97 |
| Estonia | 1322920 | 69 | 27 | 98740.406 |
| Sweden | 9971638 | 87 | 23 | 107819.278 |
| Norway | 5337962 | 82 | 17 | 24116.983 |
| Finland | 5522576 | 85 | 16 | 17176.113 |
| Iceland | 336713 | 94 | 3 | 19208.791 |

## Results

The CC values (per 100 persons, blue “crosses”) versus the volume of population, its density and the urbanization level are shown in Figs. 1-6. We have used datasets from Tables 1 and 2. The best fitting lines (black) were calculated by the least squares method [25]. The linear regression was used to calculate the regression coefficient *r* and the coefficients *a* and *b* of corresponding straight lines, [25]:

**Fig.1.**
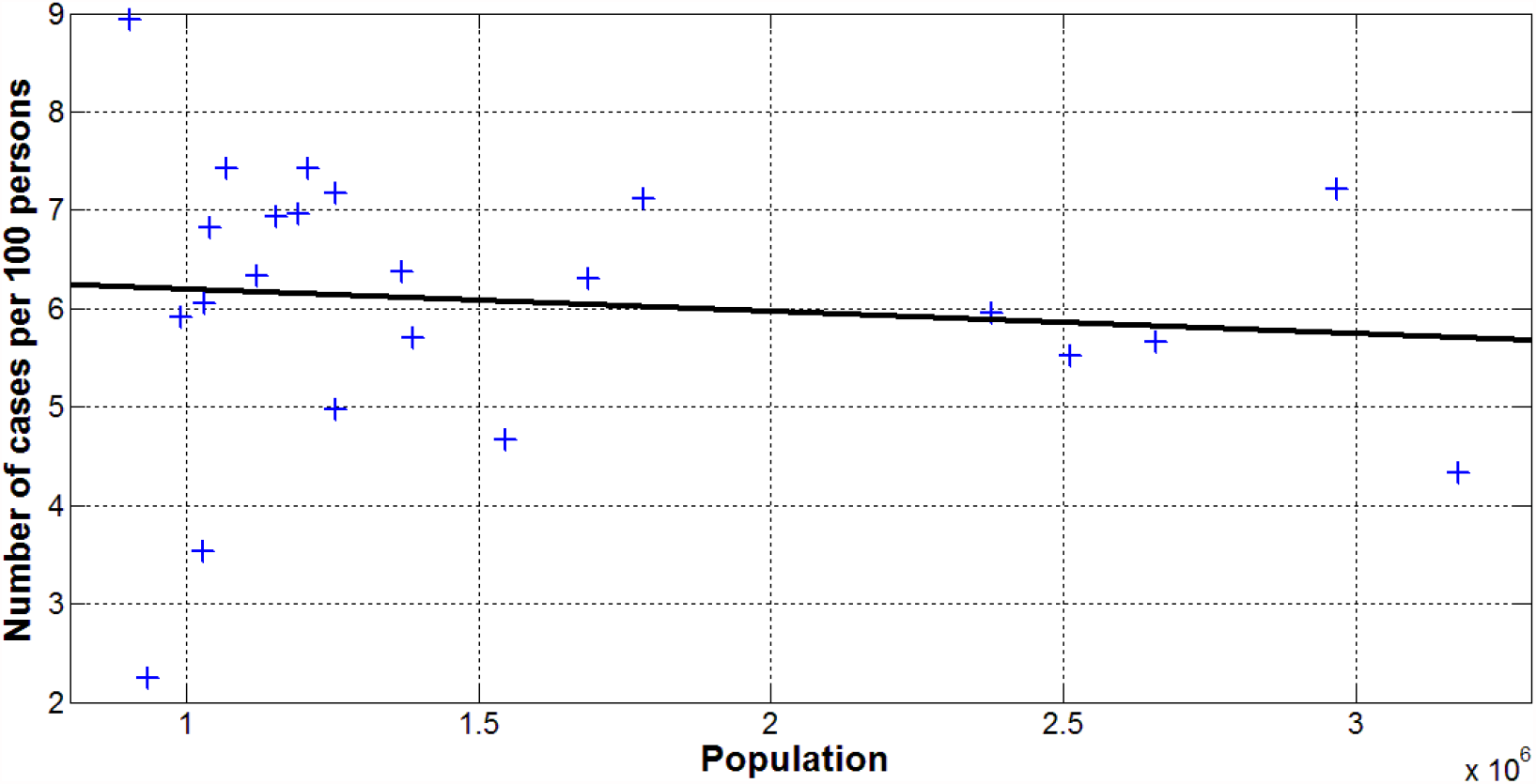
COVID-19 cases per 100 persons versus volume of population registered in the regions of Ukraine as of June 27, 2021. Blue “crosses” show the accumulated number of laboratory-confirmed cases per 100 persons (Table 1). The best fitting line (3) is shown in black.

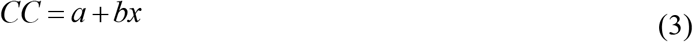

where *x* is the volume of population *N*_*pop*_ (Figs. 1 and 2), its density per square km (Figs. 3 and 4), and the urbanization level N_urb_/*N*_*pop*_ (Figs. 5 and 6).

**Fig. 2.**
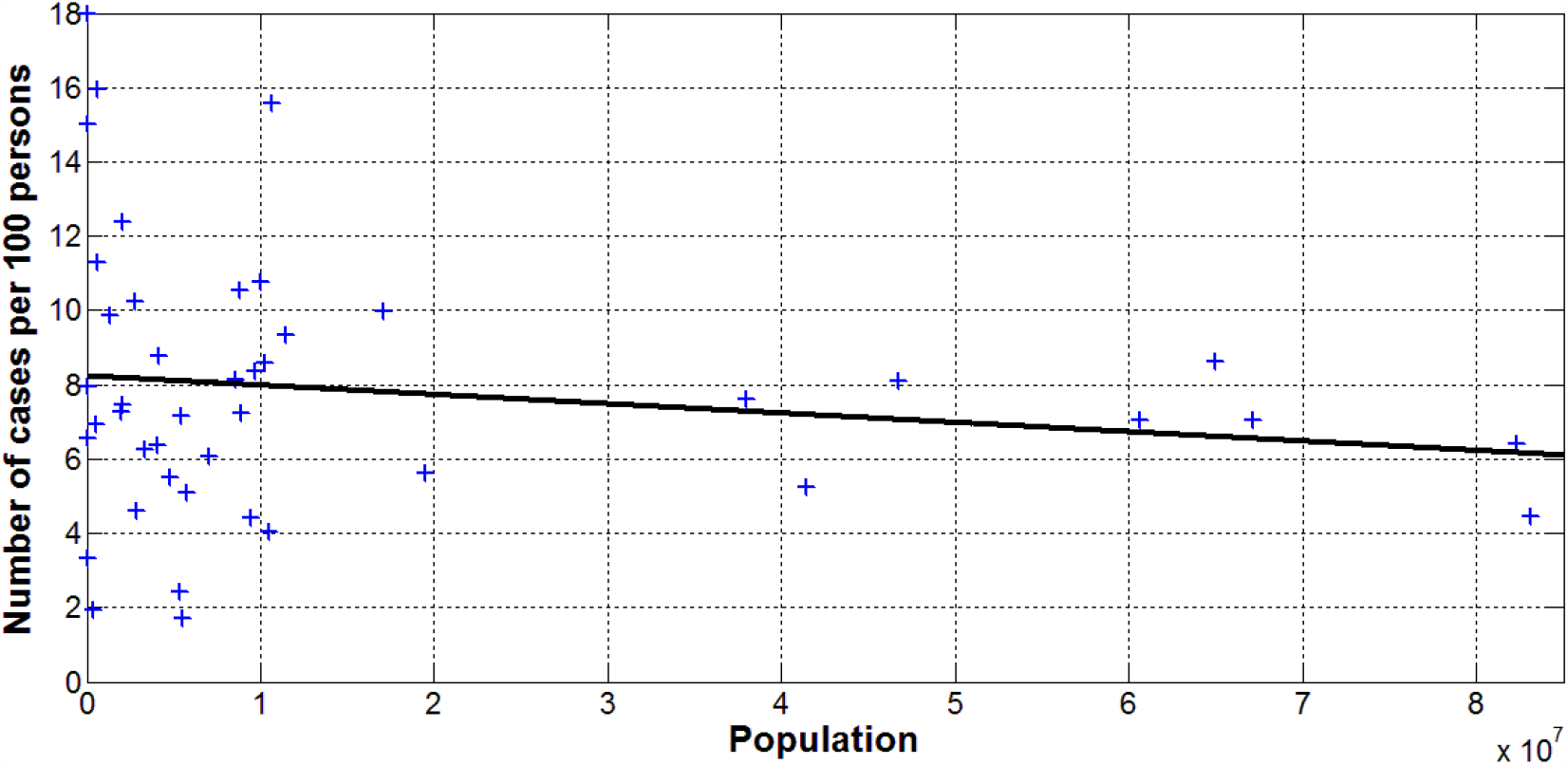
COVID-19 cases per 100 persons versus volume of population registered in the European countries as of June 28, 2021. Blue “crosses” show the accumulated number of laboratory-confirmed cases per 100 persons (Table 2). The best fitting line (3) is shown in black.

**Fig. 3.**
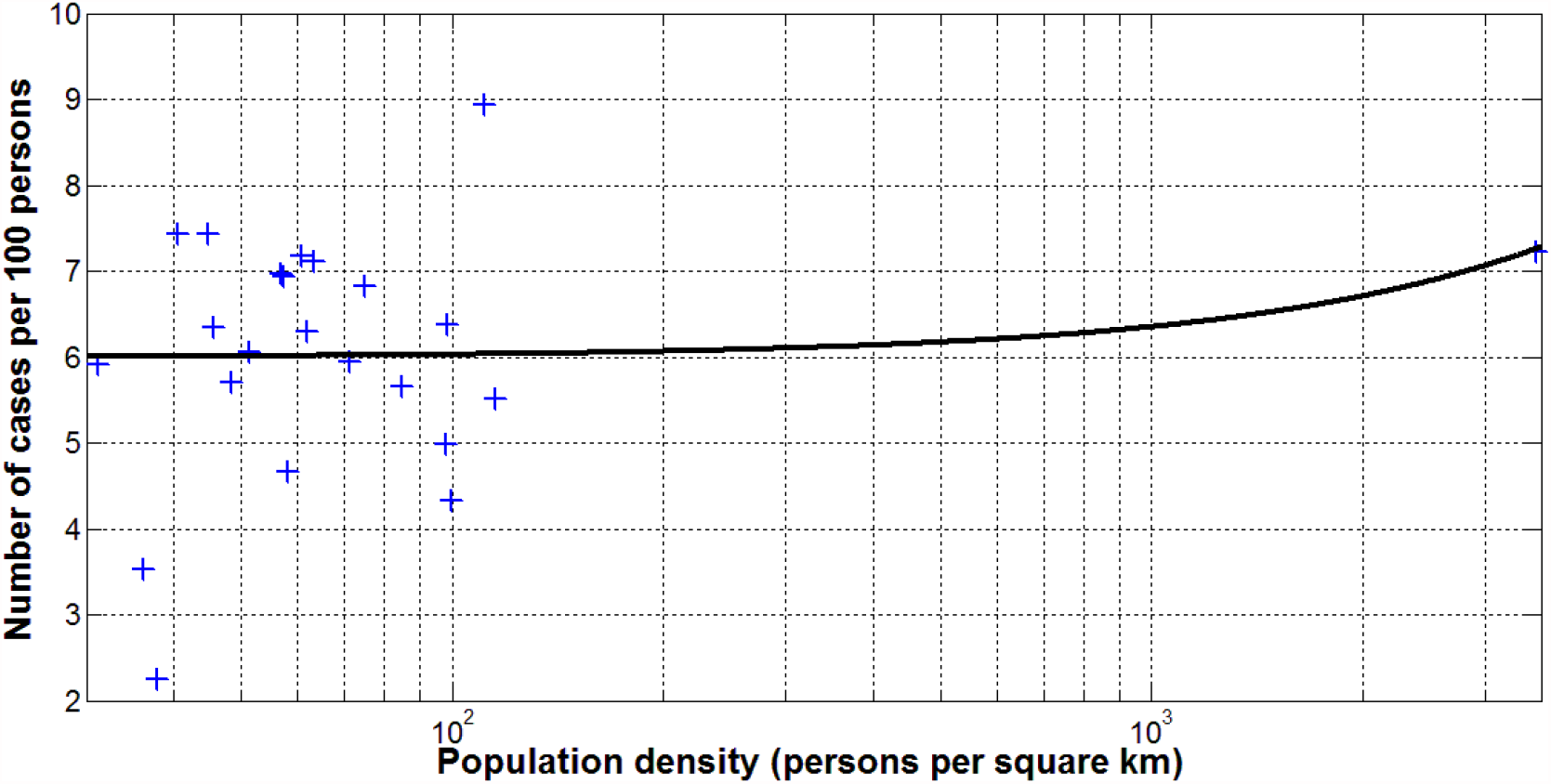
COVID-19 cases per 100 persons versus density of population (per square km) registered in the regions of Ukraine as of June 27, 2021. Blue “crosses” show the accumulated number of laboratory-confirmed cases per 100 persons (Table 1). The best fitting line (3) is shown in black.

**Fig. 4.**
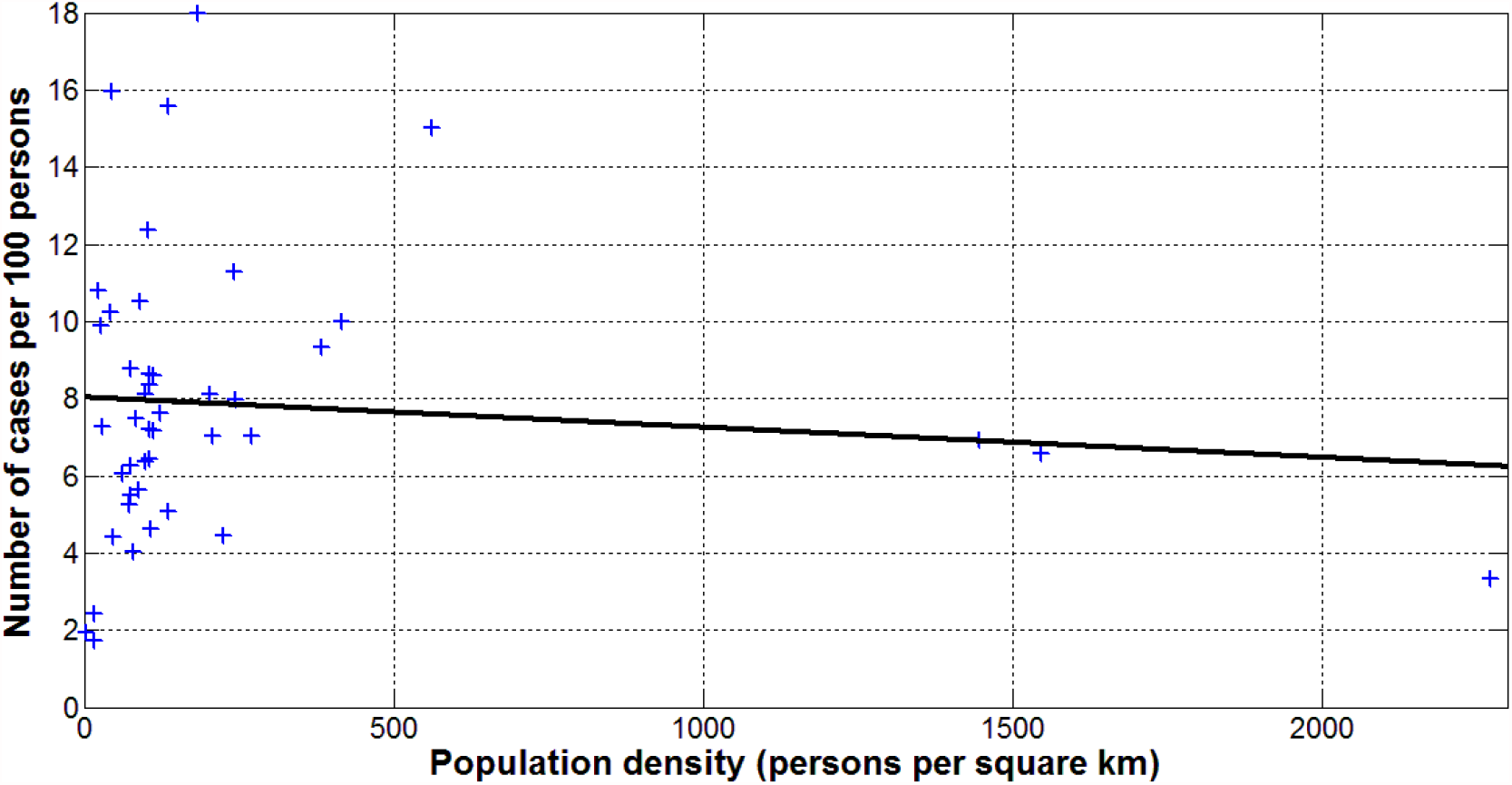
COVID-19 cases per 100 persons versus density of population (per square km) registered in the European countries as of June 28, 2021. Blue “crosses” show the accumulated number of laboratory-confirmed cases per 100 persons (Table 2). The best fitting line (3) is shown in black.

**Fig. 5.**
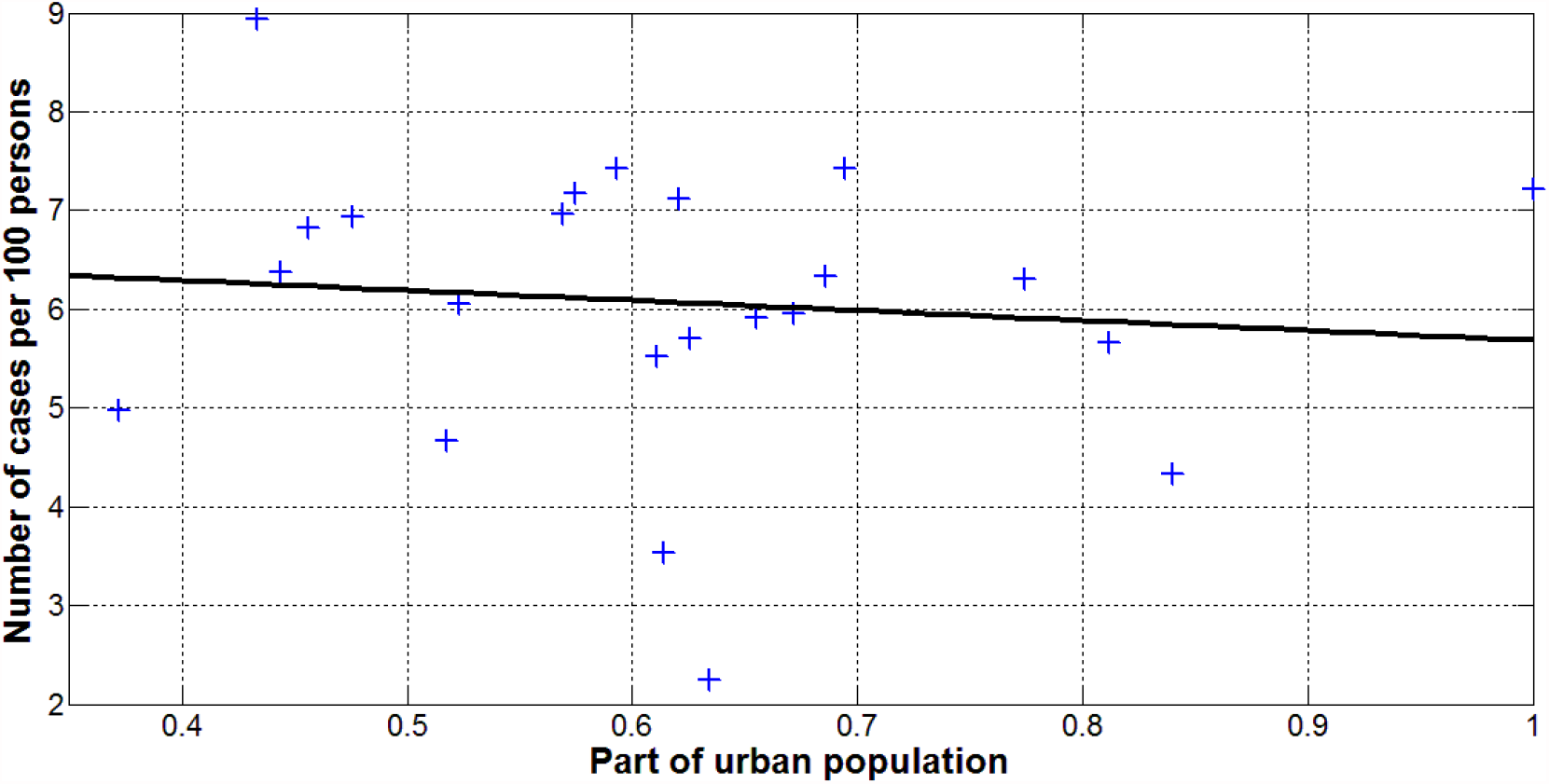
COVID-19 cases per 100 persons versus part of the urban population *N*_*ubr*_/*N*_*pop*_ registered in the regions of Ukraine as of June 27, 2021. Blue “crosses” show the accumulated number of laboratory-confirmed cases per 100 persons (Table 1). The best fitting line (3) is shown in black.

**Fig. 6.**
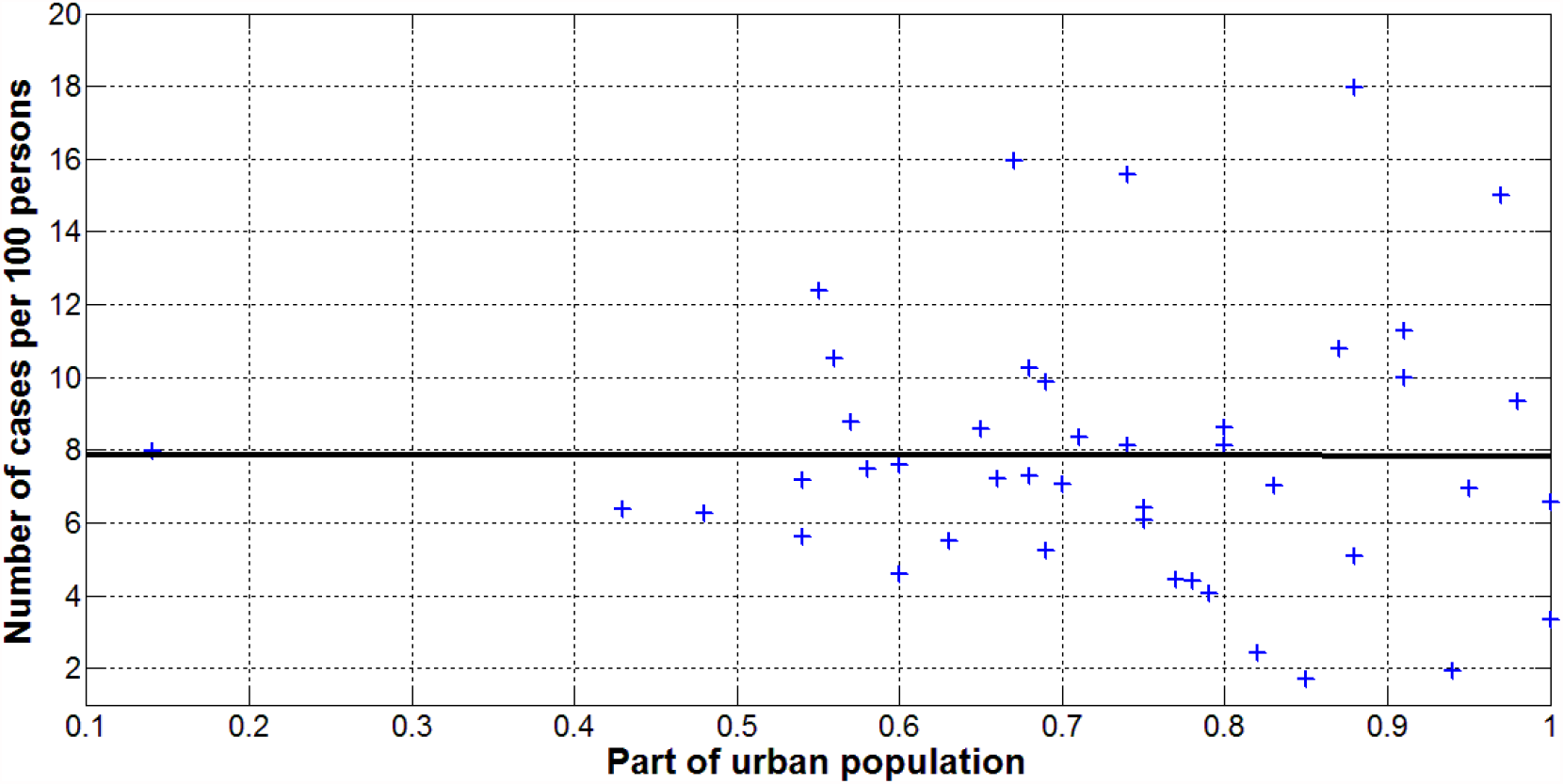
COVID-19 cases per 100 persons versus part of the urban population *N*_*ubr*_/*N*_*pop*_ population registered in the European countries as of June 28, 2021. Blue “crosses” show the accumulated number of laboratory-confirmed cases per 100 persons (Table 2). The best fitting line (3) is shown in black.

Figs. 1 and 2 illustrate that there is no visible correlation between CC values and the volume of population both in the case of Ukrainian regions and European countries. We can see only slight decreasing of CC values with increasing of *N*_*pop*_. The same tendency was revealed in [19] for the 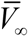 (see eq. (1)). Corresponding values of *r, a, b* and the number *n* of regions or countries taken for calculations are shown in Table 3.

**Table 3.**
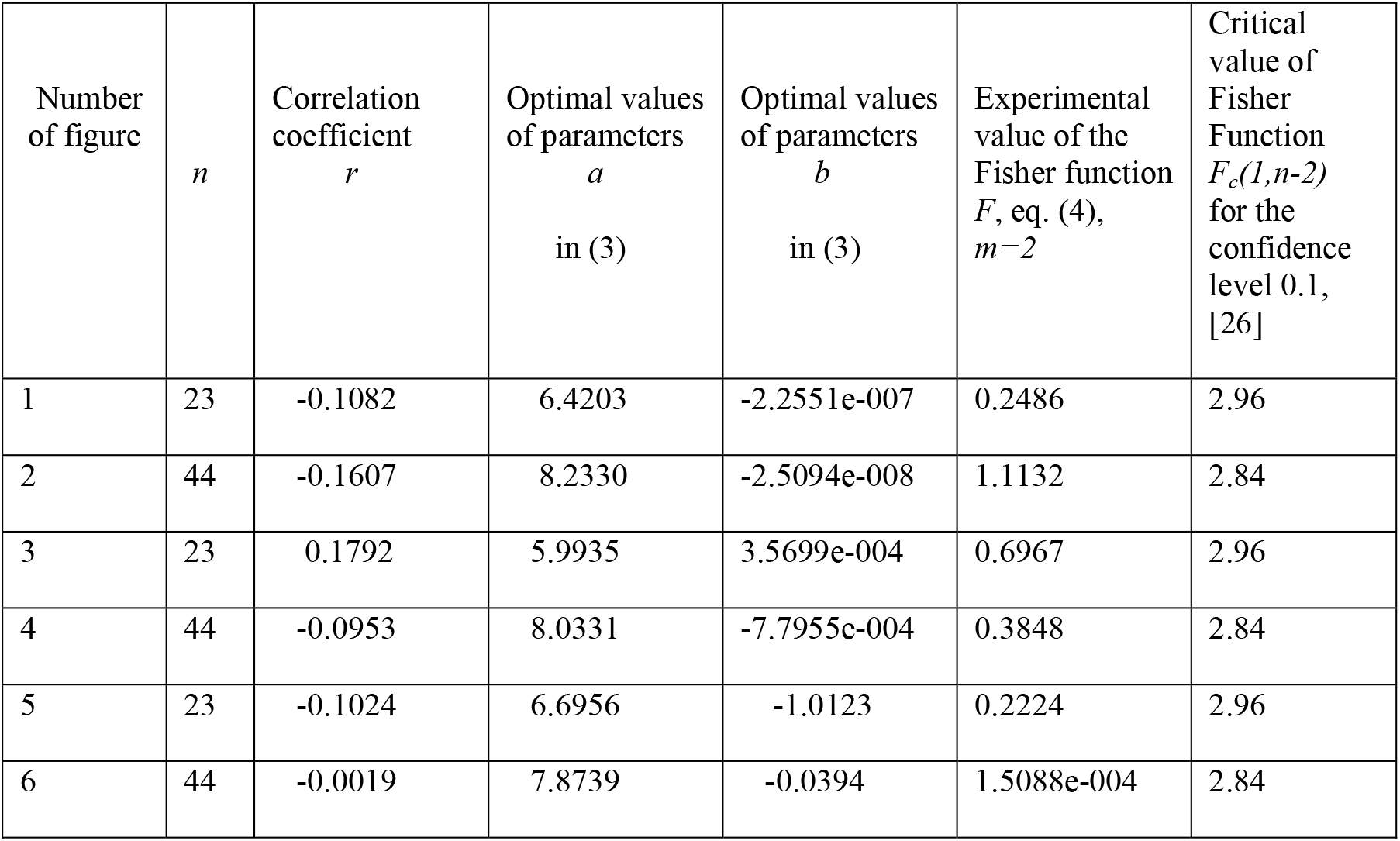
Optimal values of parameters in eq. (3), correlation coefficients and the results of Fisher test applications.

Figs. 3-6 and Table 3 illustrate that CC values do not correlate with the density of population and the urbanization level both in the case of Ukrainian regions and European countries. In Fig. 3 we can see only slight increasing of CC values with increasing of the density of population in Ukrainian regions. Opposite trend is visible in Fig. 4 for European countries. The corresponding values of the correlation coefficient and parameter *b* have opposite signs. In Figs. 5 and 6 we can see a slight decreasing of CC values with increasing the level of urbanization *N*_*ubr*_/*N*_*pop*_.

## Discussion

We can use also the F-test for the null hypothesis that says that the proposed linear relationship (3) fits the data sets. The experimental values of the Fisher function can be calculated with the use of the formula:

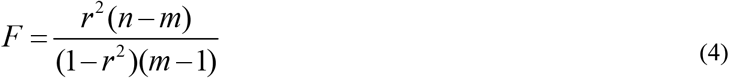

where *m*=2 is the number of parameters in the regression equation, [25]. The corresponding experimental values *F* are shown in Table 3. They have to be compared with the critical values *F*_*C*_ (*k*_1_, *k*_2_) of the Fisher function at a desired significance or confidence level (*k*_1_ = *m* −1, *k*_2_ = *n* − *m*, see, e.g., [26]). Comparisons of the values in the last two columns of Table 3 show that the critical values are much higher than the experimental *F* values. It means that the data sets presented in Tables 1 and 2 do not support the linear relationship (3).

We have checked also the non-linear dependences:

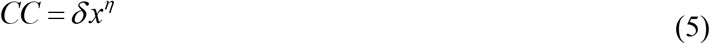

instead of (3) as it was done in [19, 27, 28]. Similar to the case of linear dependence (3), the calculations showed that corresponding values |*r*| << 1 and *F*_*C*_ (*k*_1_, *k*_2_) > *F*. It means that hypothesis (5) was not also supported by the datasets presented in Tables 1 and 2.

Very different CC values registered in the regions of Ukraine and European countries could be a result of different coronavirus strains, quarantine measures, testing, tracing and isolating patients. One more reason may be the large number of unregistered cases observed in many countries [29-33]. Estimates for Ukraine and Qatar made in [29, 33] showed that the real number of cases is about 4-5 times higher than registered and reflected in the official statistics. Similar estimates can be made for the regions of Ukraine and other European countries.

If we apply the visibility coefficients *β*_10_ =3.7 and *β*_3_ = 5.308 calculated for the Ukraine and Qatar (see [29, 33]) and take accumulated numbers of laboratory-confirmed cases, the CC values could be estimated as 20% in Ukraine and 42% in Qatar (as of the end of June 2021). Such a high percentage of people who catch the coronavirus infection can significantly affect the evaluation of the vaccination efficiency. Probably this is why we do not yet see the effect of vaccination on the pandemic dynamics in Qatar, [34].

The highest CC values registered in Europe are close to ones in other regions, e.g., Seychelles-15.8%, Bahrain-15.6%. The lowest CC values in Europe is much higher than in Vanuatu, Micronesia, Tanzania (around 0.001%), and China (0.006%). Very small CC values in the WHO Western Pacific region (e.g. Vietnam – 0.017%; Laos -0.029%; South Korea, Cambodia -0.3%; and 0.63% for Japan) need special investigations, but let us express some hypotheses.

First of all the COVID-19 pandemic probably started in this region in August 2019, [18]. It means that first cases were not identified and registered during at least 4 months. Probably these first cases were not very severe and the symptoms were not so pronounced. Presumably mutations of the coronavirus made it more pathogenic and sick people became more noticeable in December 2019. But previous cases were not taken into account in the statistics. Recent DNA investigations of East Asia population reported the presence of coronavirus around 20,000 years ago [35]. Probably, the population of this region had a collective immunity to pathogens similar to Covid-19 before the pandemic.

## Data Availability

data is in the text

